# Specific concussion curriculum: Does it improve residents’ comfort, knowledge, and In-Training Examination scores? A pilot study

**DOI:** 10.1101/2021.04.17.21255666

**Authors:** Sandal Saleem, Jessica Jary, Kelly Levasseur

## Abstract

**Background:** Concussion is defined as aberrant brain function consequent to traumatic injury characterized by disorientation or loss of consciousness or memory. If concussions are not recognized and treated appropriately, they can cause significant morbidity. Because ∼20% of sports-related concussions occur in juveniles, pediatricians must be able to treat this injury. Our primary objective was to assess the comfort and competence of pediatric residents in managing patients with concussions. Secondary objective was to assess the change in the In-Training Examination (ITE) scores after instituting a novel multimodal concussion curriculum.

**Method:** From February to June 2019, all pediatric residents (N = 24) were required to complete a multimodal concussion curriculum, including board review-style questions, lectures, and rotation in a concussion clinic. Residents voluntarily participated in a pre-and post-curriculum survey. ITE scores from 2018 and 2019 were compared.

**Results:** Twenty-three of 24 (96%) residents completed both pre- and post-curriculum surveys; of those, 17 (74%) had matched identifiers. Most residents (82%) want more education about concussion management. Residents’ scores on knowledge-based questions increased an average of 0.64 questions, with PGY-1 showing the most improvement. The proportion of residents who correctly answered the ITE head injury/concussion questions increased from 0.33 to 0.88. The concussion clinic was identified as the most helpful tool.

**Conclusion:** To better educate pediatric residents about concussions, we propose a unique multimodal curriculum. We found improved self-assessed comfort and performance on knowledge-based questions and the ITE. We recommend that pediatric and other residency training programs consider implementing this type of curriculum.

## BACKGROUND

About 1.7 million reported concussions occur per year in the United States, 20% of which involve children and adolescents with sports-related injuries.^1,2^ A surveillance report estimated that each year between 2010 and 2016, approximately 283,000 patients under age 18 were treated in the emergency department for concussions.^3^ Mild traumatic brain injury (TBI), commonly referred to as concussion, can be defined as aberrant brain function consequent to traumatic injury that is characterized by disorientation or loss of consciousness or memory.^4,5^ It is a clinical diagnosis. Although it has been widely believed that concussions occurred only in professional athletes, evidence has shown that TBIs are more common than most pediatricians realized and are an important cause of morbidity among pediatric patients.^5,6^ Proper management of concussion is critical to reduce the long-term sequelae, especially because concussions are not completely preventable.^7,8^ Sequelae, including post-concussion syndrome and chronic neurologic disease, can be devastating because they affect activities of daily life, including school performance and interpersonal relationships.^7,9^ To promote safety in youth sports, all 50 states in the U.S. have passed concussion legislation requiring mandatory removal from play; mandatory bench time; medical clearance for return to play; training for coaches, parents, and athletes; and informed consent of parents and athletes^10,11^ This mandatory legislation with state-based variations accounted for an increase in the number of pediatric patients presenting to emergency departments and primary care offices for concussion checks and clearance before returning to play.^1,11^

Physicians in the emergency department as well as pediatricians must be able to correctly diagnose and manage concussions in pediatric patients. However, physicians’ knowledge of concussion is lacking, in part because residents from different sub-specialties might not receive adequate education about concussions.6,9,12 A study from *Pediatrics* showed that 24% of residency programs do not provide education about concussion management in their curriculum.13 The American Board of Pediatrics In-Training Examination (ITE), a yearly examination required of all pediatric residents in the U.S., indicated that the pediatric residents at Beaumont Children’s Hospital did poorly on questions related to concussion. Because our pediatric residency program did not provide formal training on diagnosing and treating concussions, those results catalyzed us to create a concussion curriculum. In other residency programs, a structured, multimodal curriculum, including attendance at a weekly morbidity and mortality conference, problem-based learning conferences, and focused reading assignments, was shown to increase ITE scores.14,15 Hence, we developed a multimodal concussion curriculum that included lectures, board-style questions, and rotation at a concussion clinic.

Our primary aim was to assess the comfort and competence of pediatric residents in managing patients with concussion. Our secondary objective was to assess the change in ITE scores after instituting the multimodal concussion curriculum.

## METHOD

We conducted this study at a large suburban teaching hospital in the Beaumont Health System, where there are over 90 residency and fellowship programs, including a pediatric residency program and an internal medicine-pediatrics residency program. There are 8 pediatric residents per postgraduate year (PGY).

This study was approved by our Institutional Review Board. All 24 pediatric residents at our institution were required to complete a concussion curriculum from February to June 2019. This curriculum consisted of (a) an hour-long Centers for Disease Control and Prevention (CDC) approved online certificate course on concussion for physicians, including mandatory completion of end-of-course questions; (b) 10 board review–style questions completed in small group sessions; (c) a half-hour video-recorded lecture on return-to-play and current AAP guidelines; and (d) one half-day concussion clinic rotation where residents were actively involved in assessing and managing 3-5 patients with concussion.

All residents were asked to participate voluntarily in pre- and post-curriculum surveys sent electronically via Qualtrics. All pediatric residents received a survey before the start of the curriculum and a different one upon completion of the curriculum. Residents were excluded if they declined to participate in the survey or if they completed only the pretest or posttest survey but not both. Surveys were anonymous, but all participants were asked to identify their PGY. The pre-and post-curriculum surveys were matched using a unique identifier, their mother’s date of birth. The pre-curriculum survey had a total of 19 questions—10 questions that assessed the resident’s comfort and experience in diagnosing and treating patients with concussions and 9 board-style knowledge-based questions. The post-curriculum survey also had a total of 19 questions. Four of the 10 comfort questions and all 9 board-style questions were repeated. The remaining 6 questions pertained to the concussion curriculum. The 9 board-style, knowledge-based questions were adapted from the study conducted by Boggild and Tator.16 These 9 multiple choice questions were scored either 0 (incorrect) or 1 (correct). Four of the 9 knowledge-based questions provided the option to check all that apply. These were scored 1 point if all correct options were selected.

The ITE was taken by all pediatric residents. The Pediatric Residency Program Director receives individual resident and class scores. The ITE scores evaluated for this study were aggregated by class (PGY-1, -2, and -3) and were reported as the proportion of residents answering each category of questions correctly. We reviewed the concussion/head injury questions under the emergency medicine section of the ITE for 24 residents who took the ITE in 2018 and 16 residents who took the ITE in 2019. PGY-3 from 2018 did not take the ITE in 2019 because they had graduated. These scores, along with the scores on the knowledge-based questions in the survey, were used to determine whether the curriculum successfully improved the residents’ understanding of how to diagnose and treat concussion.

To assess the comfort level of the pediatric residents in treating children with concussion, we compared Likert-scale scores from the pre- and post-curriculum surveys. The scale was numbered from 5 to 1 (strongly agree to strongly disagree) for the purpose of analysis. Univariate and bivariate frequency tables with associated percentages were used to summarize study findings.

## RESULTS

Twenty-three of 24 (96%) residents completed both pre- and post-curriculum surveys; of those, 17 of 23 residents (74%) had matched unique identifier and were included in the study (Table 1). Most residents (76.4%) had cared for ≤5 patients with a concussion in the past 6 months of their residency. In the pre-curriculum survey, 14 (82%) indicated that they needed more education about concussions. Only one resident had previously completed a formal course in diagnosis and management of concussion.

Prior to taking the curriculum 10 out of 17 (59%) felt uncomfortable with their concussion management knowledge. This decreased to 2 out of 17 (12%) post curriculum. Out of the 3 self-assessed comfort questions, residents reported the greatest positive change in anticipatory guidance of symptoms (Figure 1).

**Figure 1:**
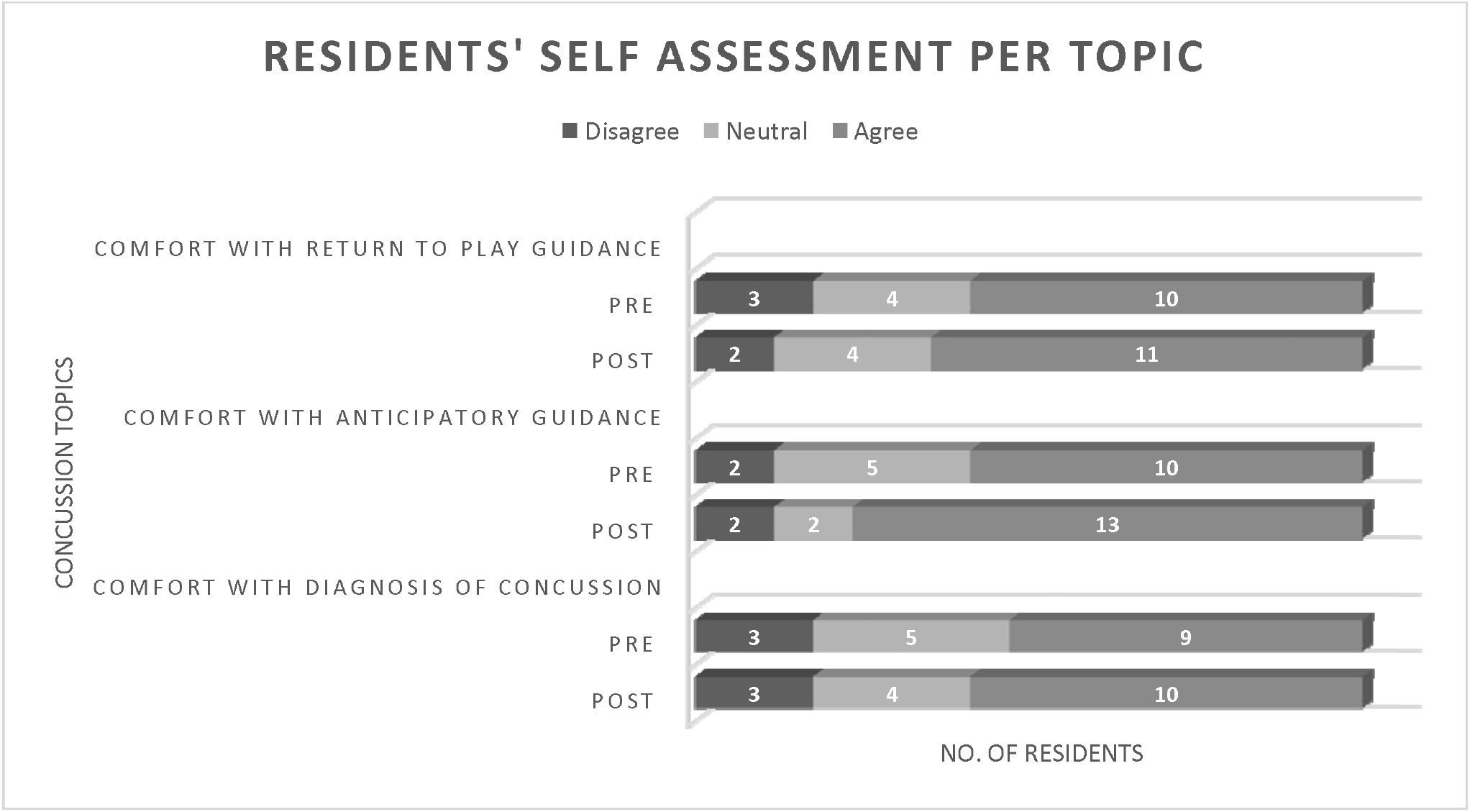
Change in resident self-assessment per concussion topic before and after the curriculum.

On average, residents scored 0.64 questions better on the post-versus pre-curriculum knowledge-based questions. 5 residents scored worse on the knowledge-based questions, 3 had no change, and 9 improved their score. First-year pediatric residents had the most improved score (Figure 2). The question that had the biggest increase in correct answers in the post-curriculum survey was “what are some ‘red flags’ that may predict the potential for more prolonged symptoms and may influence your investigation and management of concussion?”

**Figure 2:**
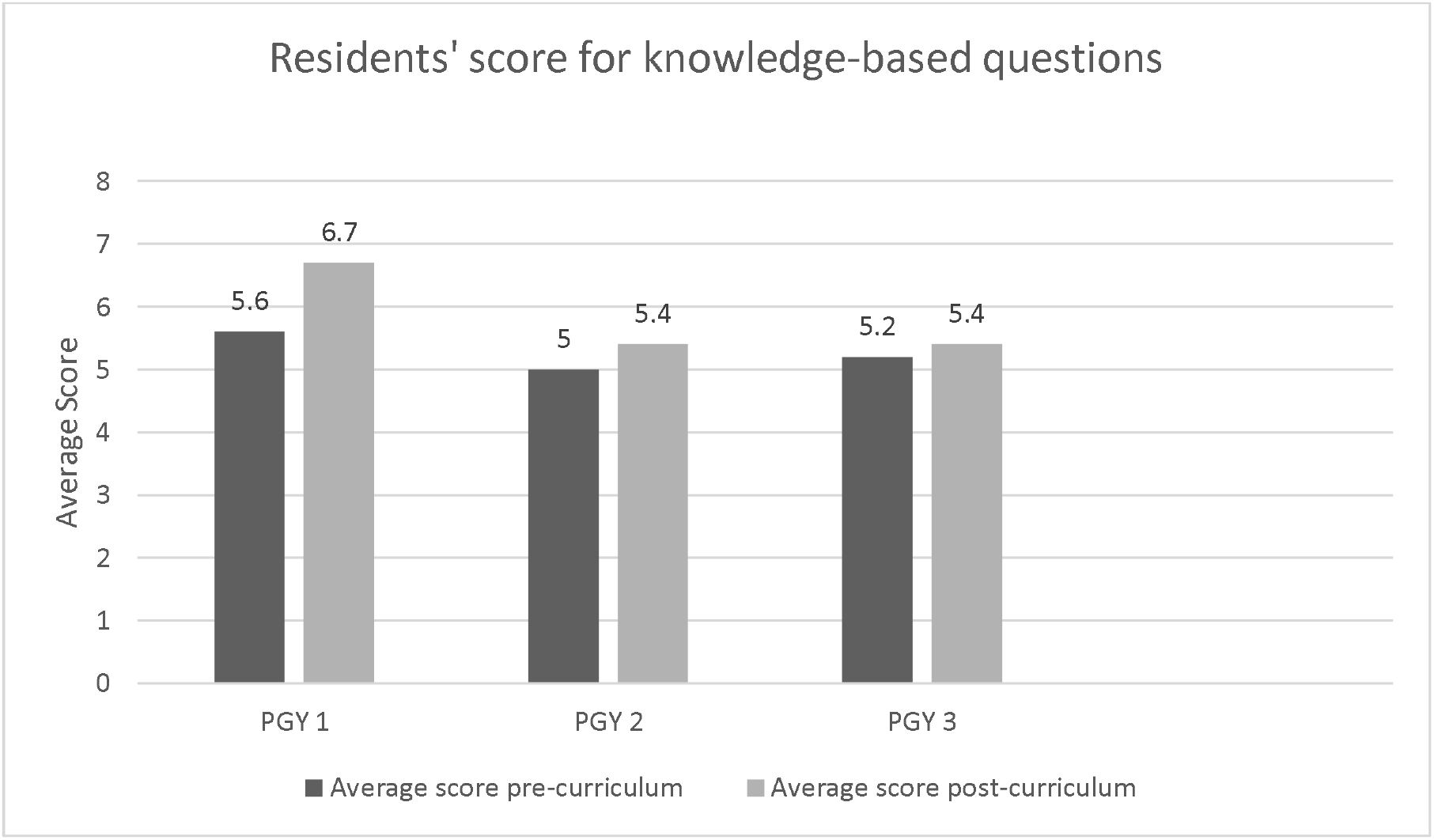
Change in residents’ score for knowledge-based questions before and after the curriculum.

The ITE scores for PGY-1, -2, and -3 were compared for 2018 and 2019. The proportions of trainees from each class who answered the concussion/head injury questions correctly on the 2018 ITE were 0.22, 0.50, and 0.29, respectively. In 2019, only data for the previous PGY-1 and PGY-2 classes were available because the PGY-3 class had graduated. The proportions of trainees from the PGY-1 and PGY-2 classes who answered the questions correctly were 0.80 and 1.00, indicating that 80% and 100% of the respective classes answered those questions accurately.

To evaluate the utility of each component of the curriculum, the post-curriculum survey asked the participants to identify the most helpful educational format. They indicated concussion clinic (47%) followed by board review-style questions (29%) and online training (24%). Residents indicated that lectures (47%) were preferred for future education. Overall, 41.2% (7/17) of the residents recommended keeping this curriculum for future residents; PGY 1 (4/7 or 57%) were most in favor.

The curriculum can be implemented by different residency programs as either part of a rotation or as a longitudinal course. The concussion clinic can be arranged in house or as an outside rotation. Simulation of 2-5 cases can be arranged as an alternative to the concussion clinic during the curriculum. The online curriculum material is available free, allowing for easy access for residents during their training.

## DISCUSSION

The number of pediatric patients with concussion has been increasing, making it important for physicians to be able to diagnose and treat these patients effectively. Currently, residency programs do not have or do not emphasize specific concussion curricula; therefore, physicians are graduating without optimal knowledge to adeptly evaluate and manage concussions.6,12,13,17 Not all educational curricula increase physicians’ comfort and knowledge equally. One previous study compared improvement in knowledge between pediatric residents who were assigned to read didactic material and sports medicine residents who were assigned to rotate in a concussion clinic. Improvement in knowledge scores increased by 1.2% for the pediatric residents versus 14.4% for the sports medicine residents. Those results show that different teaching modalities can improve knowledge, but the outcome can vary considerably.17 Keeping that in mind, we developed a multimodal concussion curriculum.

Our approach, which combines different formats to accommodate different learning styles, has been substantiated by earlier studies.17,18 Estai and Bunt reviewed multiple teaching methods for an anatomy curriculum and evaluated their effectiveness from the perspectives of students, clinicians, and anatomists. They concluded that a multimodal curriculum, including prosection, medical imaging, living anatomy, and computer-based learning, was the best teaching methodology for medical students; it also received the greatest support from the anatomists.18

We found that residents’ self-assessed comfort with diagnosis, anticipatory guidance regarding symptoms, and return-to-play guidelines improved after the curriculum. Their knowledge, based on scores from knowledge-based questions in the survey and the proportion of residents answering concussion questions correctly on the ITE, also improved after the curriculum. These findings are supported by studies that assessed ITE scores of otolaryngology and surgery residents after implementing a curriculum.14,15

Our study showed that rotation in a concussion clinic is an important component of resident learning. The survey found that most residents, regardless of PGY level, had seen ≤5 patients with concussion. Because they might not have the opportunity to care for patients with concussions regularly in their residency clinics, they need training in evaluating and managing these patients in a different setting, such as a dedicated concussion clinic. We propose that, if residents are more comfortable caring for patients with concussions, this could result in more treatment in pediatric clinics, which would decrease the need for consultation by subspecialists, specifically neurologists, who have seen a steady increase in concussions (36% increase in 2008–2009, 84% in 2009–2010, and 150% in 2011–2012).11,19

Executing the curriculum over a year would give the residents more time to complete the curriculum. The use of structured curriculum development and outcome assessment framework would give an opportunity to assess the utility of each educational tool. It would have been helpful to know at 6 months and 1-year post curriculum if they retained the knowledge and, if not, determine what other changes to the curriculum could be made to help retain knowledge.

Through this study, we hope to inform our Pediatric Residency Program Directors as well as other residency directors regarding effective tools to improve resident education about concussions. We incorporated teaching about concussions via lecture and concussion clinic rotations into the curriculum for current pediatric residents at Beaumont Health System. All four components of the curriculum were included in the adolescent medicine rotation beginning with the new resident class (PGY-1) in July 2020, as we found that PGY-1 residents showed the most improvement in knowledge and recommended continuation of the curriculum, suggesting their greater need for education and greater interest in the curriculum. We had several limitations in the study. 23 of the 24 residents responded to the survey; however, we excluded 6 because their unique identifier was entered incorrectly and hence surveys could not be matched. Self-report is an inherent limitation of any survey study, but we also received specific feedback that the residents who did answer the questions might not have answered them honestly, possibly because the residents were unhappy that the concussion curriculum was “extra work.” Another limitation was the fact that a Grand Rounds lecture on concussion was not part of the curriculum and was not attended by all residents. To mitigate this limitation, we included a question in the post-curriculum survey to determine how many residents attended it and whether they thought the Grand Rounds lecture had the same educational value as the CDC- and AAP-approved online concussion course. Of those who attended Grand Rounds (11/17; 65%), 5/11 (45.5%) reported that it had the same educational value as the CDC- and AAP-approved online concussion course.

## CONCLUSION

In conclusion, we demonstrated that residents need specific concussion curriculum and recommend using a multimodal approach. We found improved self-assessed comfort and performance on knowledge-based questions and the ITE. More studies with a bigger sample size are needed to strengthen this recommendation.

## TABLES

Table 1: Resident demographics (N = 17).

## Data Availability

All data is saved with Beaumont Health repository SharePoint.

## Declarations

- Ethics approval and consent to participate: Not Applicable
- Consent for publication: Not Applicable
- Consent to participate: Participants were provided written consent with the survey questionnaire.
- Availability of data and materials: The datasets used and survey questionnaires are available from the corresponding author on reasonable request.
- Code availability: NA
- Conflict of interests: The authors declare that they have no competing interests.
- Funding: None
- Authors’ contributions: Sandal Saleem: conception and design, acquisition of data, analysis and interpretation of data drafting the manuscript, gave final approval of the version to be submitted and any revised version. Jessica Jary: conception and design, acquisition of data and gave final approval of the version to be submitted and any revised version. Kelly Levasseur: Drafting the manuscript, critical editing and gave final approval of the version to be submitted and any revised version.
- Authors’ information: Sandal Saleem MD, Phone: 501-516-0635, Role at the time of the study: Pediatric Emergency Medicine fellow Jessica Jary DO, Phone: 248-982-7406, Role at the time of the study: 3^rd^ year Pediatric resident Kelly Levasseur DO, Phone: 248-284-3270, Role at the time of the study: Pediatric Emergency Medicine Fellowship program director

## Acknowledgements

I (S.S.) thank Dr. Kelly Doyle, Pediatric Residency Program Director, for her support in implementing the curriculum and reviewing the manuscript. I thank Ms. Susan Musto, Director of the Concussion Clinic, for sharing her lecture on “Return to Play and Learning Guidelines” and for accommodating pediatric residents in the concussion clinic.

## Abbreviations

TBI: traumatic brain injury
ABP ITE: American Board of Pediatrics In-Training Examination

## REFERENCES

1. Kosoy J, Feinstein R. Evaluation and management of concussion in young athletes. Curr Probl Pediatr Adolesc Health Care. 2018;48(5-6):139–150. doi: S1538-5442(18)30050-6 [pii].

2. Pfister T, Pfister K, Hagel B, Ghali WA, Ronksley PE. The incidence of concussion in youth sports: A systematic review and meta-analysis. Br J Sports Med. 2016;50(5):292–297. doi: 10.1136/bjsports-2015-094978 [doi].

3. Sarmiento K, Thomas KE, Daugherty J, et al. Emergency department visits for sports-and recreation-related traumatic brain injuries among children - united states, 2010-2016. MMWR Morb Mortal Wkly Rep. 2019;68(10):237–242. doi: 10.15585/mmwr.mm6810a2 [doi].

4. National Center for Injury Prevention and Control. Report to congress on mild traumatic brain injury in the united states: Steps to prevent a serious public health problem. Centers for Disease Control and Prevention Web site. www.cdc.gov/traumaticbraininjury/pdf/mtbireport-a.pdf. Updated 2003. Accessed 2/10/, 2020.

5. Traumatic brian injury and concussion. Centers for Dissease Control and Prevention Web site. https://www.cdc.gov/traumaticbraininjury/get_the_facts.html. Accessed 2/10/, 2020.

6. Mann A, Tator CH, Carson JD. Concussion diagnosis and management: Knowledge and attitudes of family medicine residents. Can Fam Physician. 2017;63(6):460–466. doi: 63/6/460 [pii].

7. Rotter J, Kamat D. Concussion in children. Pediatr Ann. 2019;48(4):e182–e185. doi: 10.3928/19382359-20190326-01 [doi].

8. Sone JY, Kondziolka D, Huang JH, Samadani U. Helmet efficacy against concussion and traumatic brain injury: A review. J Neurosurg. 2017;126(3):768–781. doi: 10.3171/2016.2.JNS151972 [doi].

9. Harmon KG, Drezner JA, Gammons M, et al. American medical society for sports medicine position statement: Concussion in sport. Br J Sports Med. 2013;47(1):15–26. doi: 10.1136/bjsports-2012-091941 [doi].

10. National Center for injury prevention and control. Implementing return to play: Learning from the experiences of early implementers. Centers for Diseaase Control Web site. www.cdc.gov/headsup/pdfs/policy/RTP_Implementation-a.pdf. Accessed 2/10/, 2020.

11. Rose SC, Weber KD, Collen JB, Heyer GL. The diagnosis and management of concussion in children and adolescents. Pediatr Neurol. 2015;53(2):108–118. doi: 10.1016/j.pediatrneurol.2015.04.003 [doi].

12. Itriyeva K, Feinstein R, Carmine L. Pediatric providers’ attitudes and practices regarding concussion diagnosis and management. Int J Adolesc Med Health. 2017;31(6):10.1515/ijamh-0070. doi: 10.1515/ijamh-2017-0070 [doi].

13. Demorest RA, Bernhardt DT, Best TM, Landry GL. Pediatric residency education: Is sports medicine getting its fair share? Pediatrics. 2005;115(1):28–33. doi: 115/1/28 [pii].

14. Redmann AJ, Tawfik KO, Myer CM,3rd. The impact of a resident-run review curriculum and USMLE scores on the otolaryngology in-service exam. Int J Pediatr Otorhinolaryngol. 2018;104:25–28. doi: S0165-5876(17)30508-6 [pii].

15. Decoteau MA, Rivera L, Umali K, Chan AD, Soballe P, Ignacio RC. A multimodal approach improves american board of surgery in-training examination scores. Am J Surg. 2018;215(2):315–321. doi: S0002-9610(17)30624-4 [pii].

16. Boggild M, Tator CH. Concussion knowledge among medical students and neurology/neurosurgery residents. Can J Neurol Sci. 2012;39(3):361–368. doi: WN17738736M853L1 [pii].

17. Haider MN, Leddy JJ, Baker JG, et al. Concussion management knowledge among residents and students and how to improve it. Concussion. 2017;2(3):CNC40-0001. eCollection 2017 Nov. doi: 10.2217/cnc-2017-0001 [doi].

18. Estai M, Bunt S. Best teaching practices in anatomy education: A critical review. Ann Anat. 2016;208:151–157. doi: S0940-9602(16)30032-2 [pii].

19. Gibson TB, Herring SA, Kutcher JS, Broglio SP. Analyzing the effect of state legislation on health care utilization for children with concussion. JAMA Pediatr. 2015;169(2):163–168. doi: 10.1001/jamapediatrics.2014.2320 [doi].

